# Urinary collagen type I degradation products as common fibrosis biomarkers in chronic diseases

**DOI:** 10.64898/2026.08.26.26361420

**Authors:** Ioanna K Mina, Yaqza Hussain, Justyna Siwy, Lorenzo Catanese, Harald Rupprecht, Joachim Beige, Jan A Staessen, Jochen Metzger, Frederik Persson, Peter Rossing, Christian Delles, Joost P Schanstra, Ayman Bannaga, Antonia Vlahou, Harald Mischak, Ramesh P Arasaradnam, Agnieszka Latosinska

## Abstract

**Background:** Fibrosis, characterised by excessive accumulation of collagen type I (COL1), is a common feature of chronic diseases, including liver diseases (LDs), chronic kidney disease (CKD) and heart failure (HF). COL1 degradation products can be detected in urine by proteomics/peptidomics analyses and may serve as non-invasive biomarkers of fibrosis. We aimed to identify a common molecular signature of fibrosis across these diseases that may ultimately guide interventions to slow disease progression and prevent organ damage.

**Methods:** Using capillary electrophoresis coupled to mass spectrometry (CE-MS), naturally occurring COL1 degradation products (peptides) in the urine of patients with fibrotic disease, LDs (n=127), CKD (n=263) or HF (n=187), were investigated and compared with the same number of matched controls. Disease-associated COL1 peptides were identified separately for each condition, and peptides showing consistent associations across the three diseases were selected to define a common fibrosis signature. A support vector machine model based on the selected peptides was developed and validated in independent cohorts of patients with LDs (n=110), CKD (n=93), HF (n=32) and controls (n=643).

**Results:** We identified a common fibrotic signature consisting of 50 COL1 degradation products, mainly downregulated in fibrosis. A model based on these peptides achieved a strong performance, with an area under the receiver operating characteristic curve (AUC) of 0.935 (95% confidence interval (CI) 0.917-0.953, p<0.0001) in an external validation cohort comprising pooled disease groups (LDs, CKD, and HF) and controls. Performance was maintained in LDs, CKD and HF, with AUCs of 0.917 (95% CI 0.890-0.944, p<0.0001), 0.951 (95% CI 0.931-0.971, p<0.0001) and 0.950 (95% CI 0.903-0.997, p<0.0001), respectively. The model scores were significantly associated with fibrosis stage in LDs (p=0.0097) and with interstitial fibrosis and tubular atrophy in CKD (p=0.045).

**Conclusion:** A model of urinary COL1 peptides captures a shared collagen degradation signature across organs and diseases, enabling the non-invasive assessment of fibrosis irrespective of its origin. As these peptides exclusively reflect collagen degradation, the findings suggest impaired collagen degradation as a driver in fibrosis. Future clinical studies are warranted to evaluate the utility of this model for early fibrosis detection and earlier implementation of anti-fibrotic interventions.

## Background

Liver diseases (LDs), chronic kidney disease (CKD) and heart failure (HF) are major chronic diseases that frequently coexist, with dysfunction in one organ system contributing to disease progression in others, interactions that are increasingly recognised within the cardiovascular-kidney-metabolic syndrome^1^. Fibrosis is an important component of the progression of these diseases and represents a common pathological process that also affects other organs, including the lung and skin^2^. Fibrosis is characterized by a dysregulated wound-healing response and excessive deposition of extracellular matrix components, particularly collagen type I (COL1)^3^, resulting from an imbalance between matrix production and degradation.

Despite differences between organs and diseases, the central role of COL1 in fibrotic remodelling represents a shared biological basis that raises the possibility that therapeutic strategies targeting fibrotic processes in one organ system could be beneficial in other fibrotic diseases. Consequently, identifying a shared fibrosis molecular signature could provide a measure of ongoing fibrotic remodelling and facilitate earlier preventive and therapeutic interventions.

Specifically, previous studies have shown increased COL1 production in fibrotic liver^4^, kidney^5^, and heart^6^ tissues, primarily involving transforming growth factor-beta signalling^6–8^. In parallel, COL1 degradation is mediated by specific proteases, including matrix metalloproteinases (MMPs) and cathepsins^9,10^, which can be influenced either by changes in protease activity (for example, through regulation by tissue inhibitors of MMPs^11^) or by modifications to the COL1 itself that alter its susceptibility to proteolysis^12,13^.

COL1 consists of three chains, two collagen type I alpha 1 (COL1A1) chains and one collagen type I alpha 2 (COL1A2) chain, which are synthesised as procollagen chains consisting of an N-terminal propeptide, a central collagen domain and a C-terminal propeptide, each^14^. Following their assembly into a triple helix and the cleavage of N- and C-propeptides, mature COL1 is formed^14^. COL1 fragments are among the most abundant naturally occurring peptides in urine^15^, with the majority (>98%) being derived from mature COL1 rather than from the N- or C-terminal propeptides, indicating that they are products of collagen degradation rather than collagen synthesis. Many of these peptides have been associated with LDs^16^, CKD^17–19^ and HF^20,21^, forming a basis for peptide-based biomarker approaches. Indeed, models based on urinary peptides have already been developed for the detection of organ specific fibrosis in liver^16^ and kidney^22^, demonstrating their applicability as non-invasive biomarkers.

Fibrosis is responsible for up to 45% of deaths in the developed countries^23^, and globally approximately 1 in 4 people have liver fibrosis, 1 in 6 have kidney fibrosis, and 1 in 60 have heart fibrosis^3^. Despite this high prevalence, biopsy remains the gold standard for diagnosing fibrosis in these organs^24–27^. However, it is an invasive procedure that carries risks of complications, including, in rare cases, death^24,26^. Additionally, biopsies cannot be repeated multiple times (for the purpose of monitoring) and are prone to sampling errors^26,27^.

Collectively, we hypothesised that: 1) fibrosis may be the consequence of attenuation of COL1 degradation, and 2) a generic fibrosis signature reflecting the collagen degradation process exists in the urinary peptidome, independent of the affected organ. To test these hypotheses, we investigated the association between naturally occurring COL1 degradation products in urine and fibrotic-related chronic diseases, with the aim of developing a non-invasive, urine-based method to detect an ongoing fibrotic remodelling across organs, rather than replace organ-specific diagnostics, and ultimately support earlier treatment decisions before irreversible organ damage occurs.

## Methods

### Capillary electrophoresis coupled to mass spectrometry (CE-MS) data

Proteome analysis was performed as described previously^28^. In brief, a P/ACE MDQ CE coupled to a micro-TOF-MS was used for CE-MS analysis. The raw CE-MS data were evaluated using the MosaFinder software assigning the detected signal in silico to the list of 21559 peptides, with >5000 having information about the amino acid sequence^29^. Twenty-nine collagen fragments generally unaffected in disease were used as internal standards for normalizing peptide intensities^30^.

### Study Population

Anonymized urinary peptidome, and phenotypic data were extracted from the Human Urinary Proteome Database^29^. Due to the known associations of age^31^, sex^32^, diabetes^33,34^ and kidney function^17^ with the urinary peptides, cases were matched based on these characteristics with their respective controls. For biomarker discovery, patients with different types of LDs (**Table 1**) and confirmed liver fibrosis established by a combination of liver ultrasound, fibroscan and laboratory markers (e.g. aspartate aminotransferase to platelet ratio index), as well as liver histology in cases of uncertainty (study by Bannaga et al.^16^) were matched in a ratio 1 to 1 with control individuals from the general population studies (FLEMENGHO^35^ and Generation Scotland^36^) based on age and sex. Because all patients with LDs had an estimated glomerular filtration rate (eGFR) above 60 mL/min/1.73 m², only control individuals with eGFR>60 mL/min/1.73 m² were selected. Similarly, CKD patients with different aetiologies (**Table 1**) and interstitial fibrosis and tubular atrophy (IFTA)≥15% (biopsy-based histological assessment) were extracted from the study by Catanese et al.^22^. Because many of these patients also had type II diabetes, they were matched with individuals from the general population study (FLEMENGHO^35^) and from the type II diabetes study (PRIORITY^34^) based on age, sex and the presence of diabetes. Matching for eGFR could not be performed due to the disease-specific distribution of kidney function in the CKD cohort. Although no data from patients with confirmed heart fibrosis were available, previous studies have established the presence of heart fibrosis in the majority of patients with HF with preserved ejection fraction (HFpEF)^37,38^. Therefore, patients with HFpEF and control individuals matched for multiple parameters, including age, sex, eGFR and the presence of diabetes, were extracted from the study by He et al.^20^.

**Table 1.**
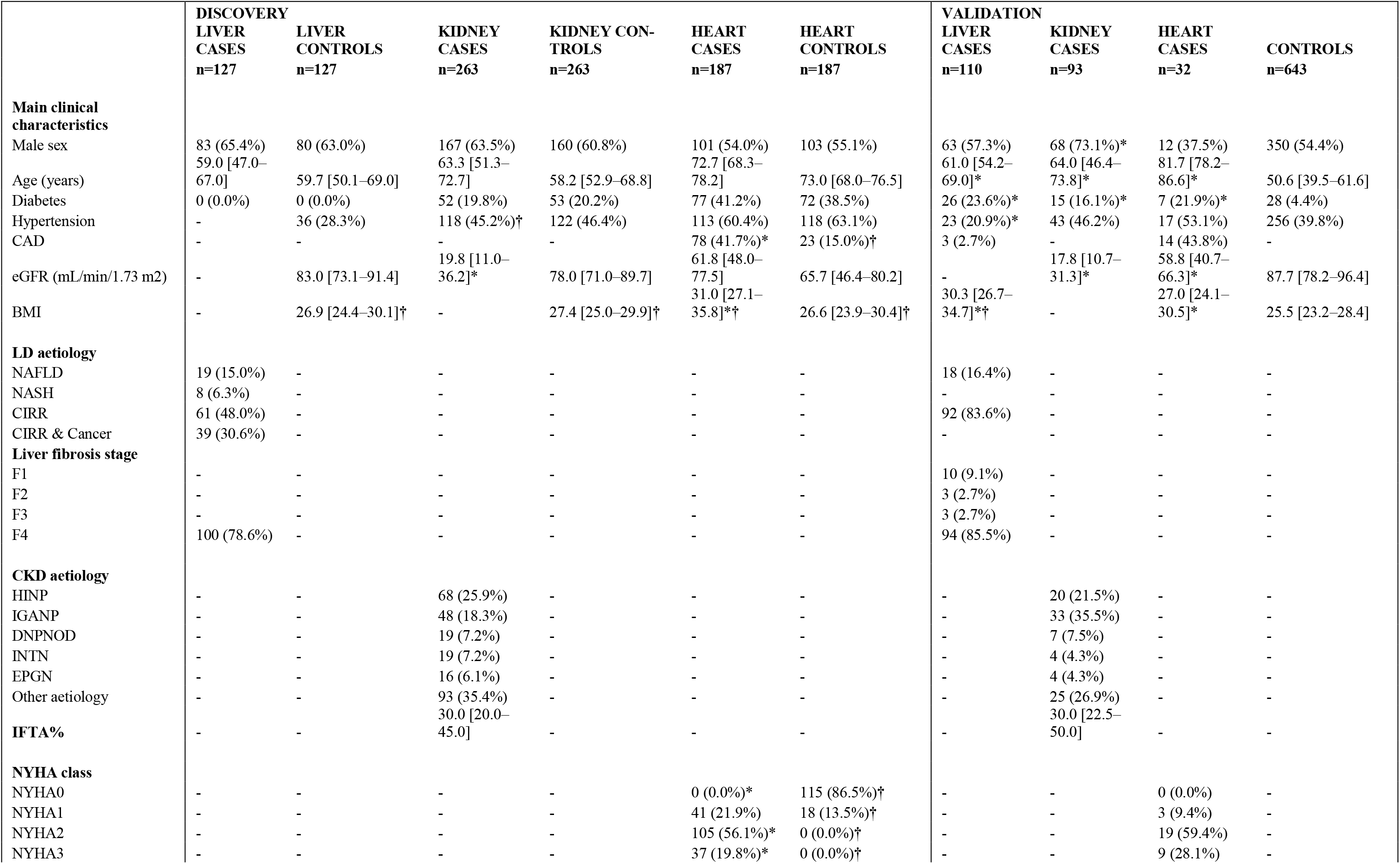
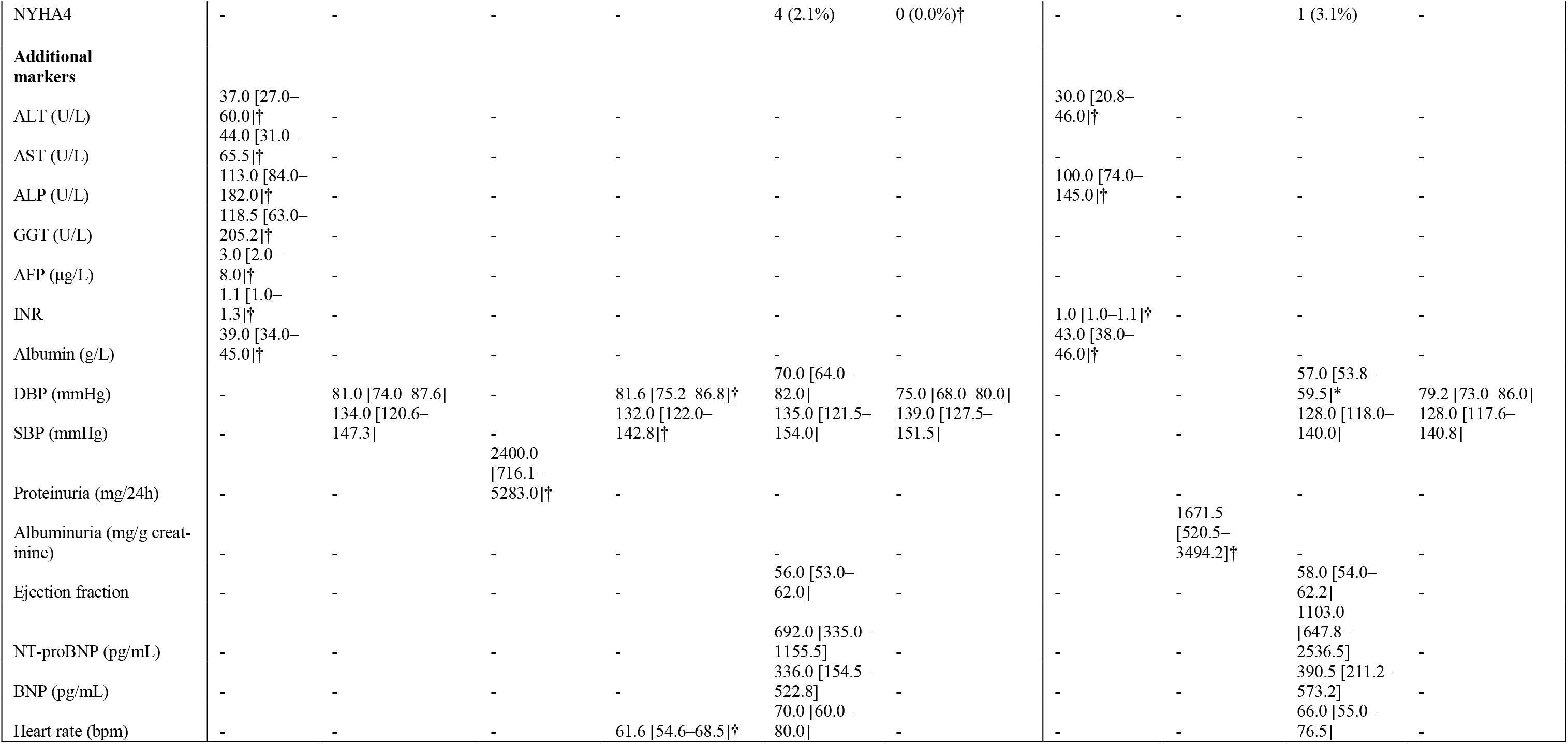
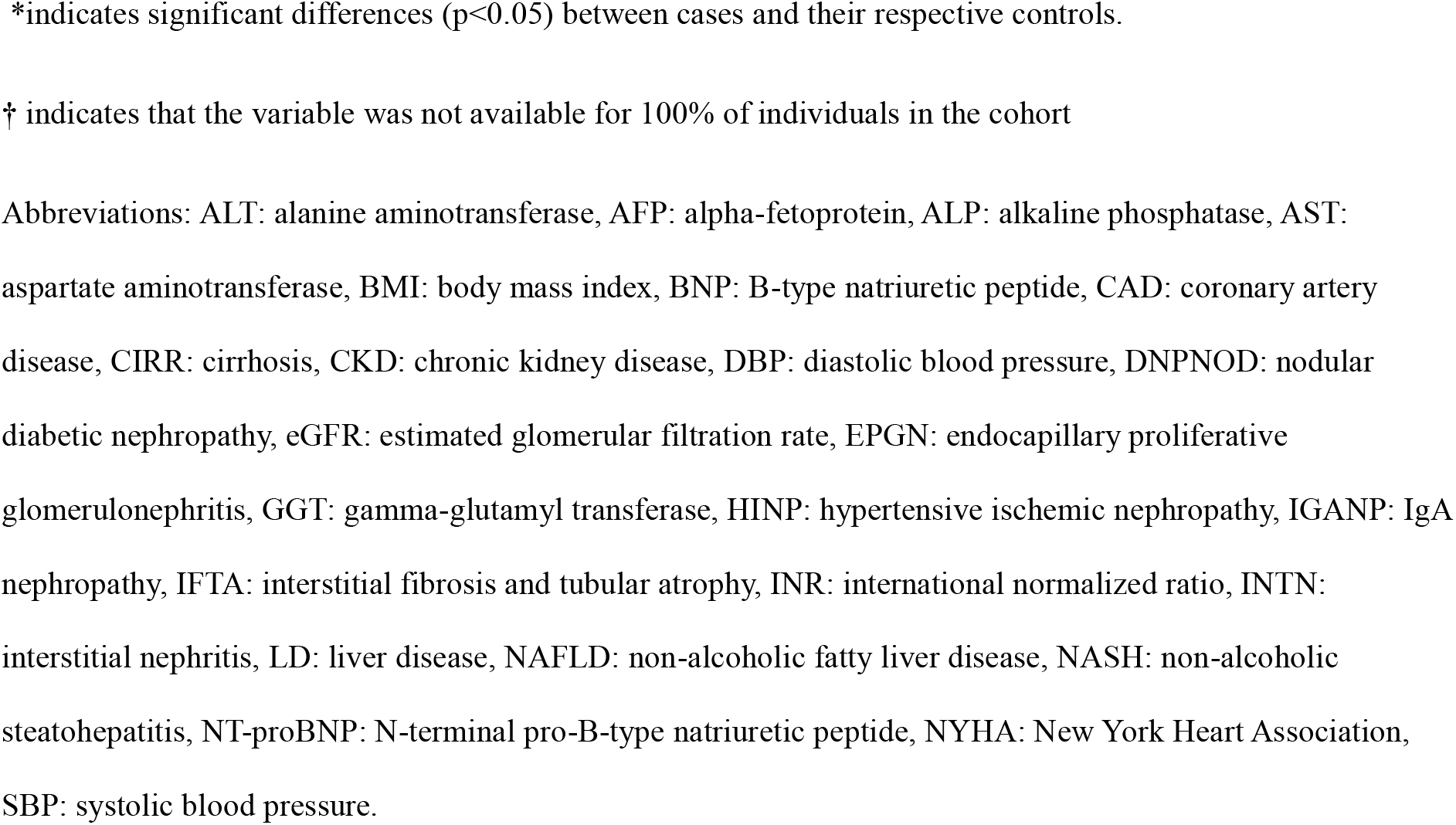
Cohort characteristics. Values are presented as median [interquartile range] for numerical variables or n (%) for categorical variables. Only characteristics with a prevalence greater than 70% in any given cohort are reported. Comparisons were made between cases and their corresponding control groups using the Mann– Whitney test for continuous variables and the Chi-squared test for categorical variables.

For the model validation independent datasets from patients with confirmed liver fibrosis (stages F1-F4), patients with confirmed IFTA≥15% and patients with HFpEF were extracted from the TENDENCY^39^ study, study by Frantzi et al.^40^ and study by He et al.^20^, respectively (**Table 1**). Liver and kidney fibrosis were assessed using the same methods applied for the discovery cohorts. For this independent validation, control individuals, who were not included in the biomarker discovery phase, without clinical signs of organ damage and with an eGFR above 60 mL/min/1.73 m², were extracted from the general population studies FLEMENGHO^35^ and Generation Scotland^36^.

Fibrosis could not be excluded in the control group (either in discovery or validation), as invasive or dedicated fibrosis assessments are not routinely performed in these populations. Only datasets from individuals above 18 years old were investigated. Cases and controls were matched using the MatchIt package in R^41^.

### Statistical analysis and model generation

Statistical analysis was performed comparing COL1 peptides (*i.e.* COL1A1, COL1A2) between cases and controls within each disease group. In each comparison, only peptides with a frequency≥30% in cases or controls were considered. The frequency threshold of 30% has been found optimal in previous peptidome studies to address missing values while maintaining high coverage^42^. Disease-associated peptides were identified using the non-parametric Mann–Whitney test and resulting p-values were adjusted using the Benjamini– Hochberg method^43^. Statistical tests were conducted under two conditions: 1) without replacing the missing values and 2) after replacing missing values with zero, and only changes with a significant adjusted p-value in the analysis after the replacement of missing values with zero, which agreed in fold change direction in the analysis without replacement of missing values, were retained. The results of the disease-specific analyses were then used for the selection of common COL1 fibrosis biomarkers. For ranking common fibrosis biomarkers based on significance, raw p-values across the three organ comparisons were combined using Fisher’s method. The analyses were conducted using the base R functions wilcox.test and p.adjust and the poolr package.

Based on selected biomarkers, a model using a support vector machine (SVM) algorithm integrated into the MosaCluster software^28^ was developed and optimised using take-one-out cross-validation. The model was trained using the same liver fibrosis patients and their respective controls that were used during biomarker development. The liver fibrosis cohort was selected for model training because fibrosis in this cohort was confirmed and eGFR was above 60 mL/min/1.73 m², reducing the potential influence of impaired kidney function on urinary peptide levels. To select the SVM hyperparameters, we screened a range of values for C (20–20,000) and gamma (0.000002–0.002). The optimal parameter combination (C: 2560; gamma 0.000002^)^ was selected based on overall cross-validation performance, with preference given to models with higher C, lower gamma, and fewer number of support vectors. The optimal model score cut-off was determined based on the cross-validation results using the Youden index. The model was then validated in independent datasets of patients with liver fibrosis, kidney fibrosis or HFpEF and individuals from the general population (**Figure 1**). The scoring of the validation cohorts based on the model was conducted in the MosaDiagnostics software^28^. The R package pROC was used to determine and plot the area under the receiver operating characteristic curve (AUC), including its confidence interval (CI), for the generated model. To assess the statistical significance of the AUC, the roc.area function from the verification package was used. Confidence intervals for sensitivity and specificity were calculated using the exact Clopper–Pearson method (https://epitools.ausvet.com.au/ciproportion).

**Figure 1.**
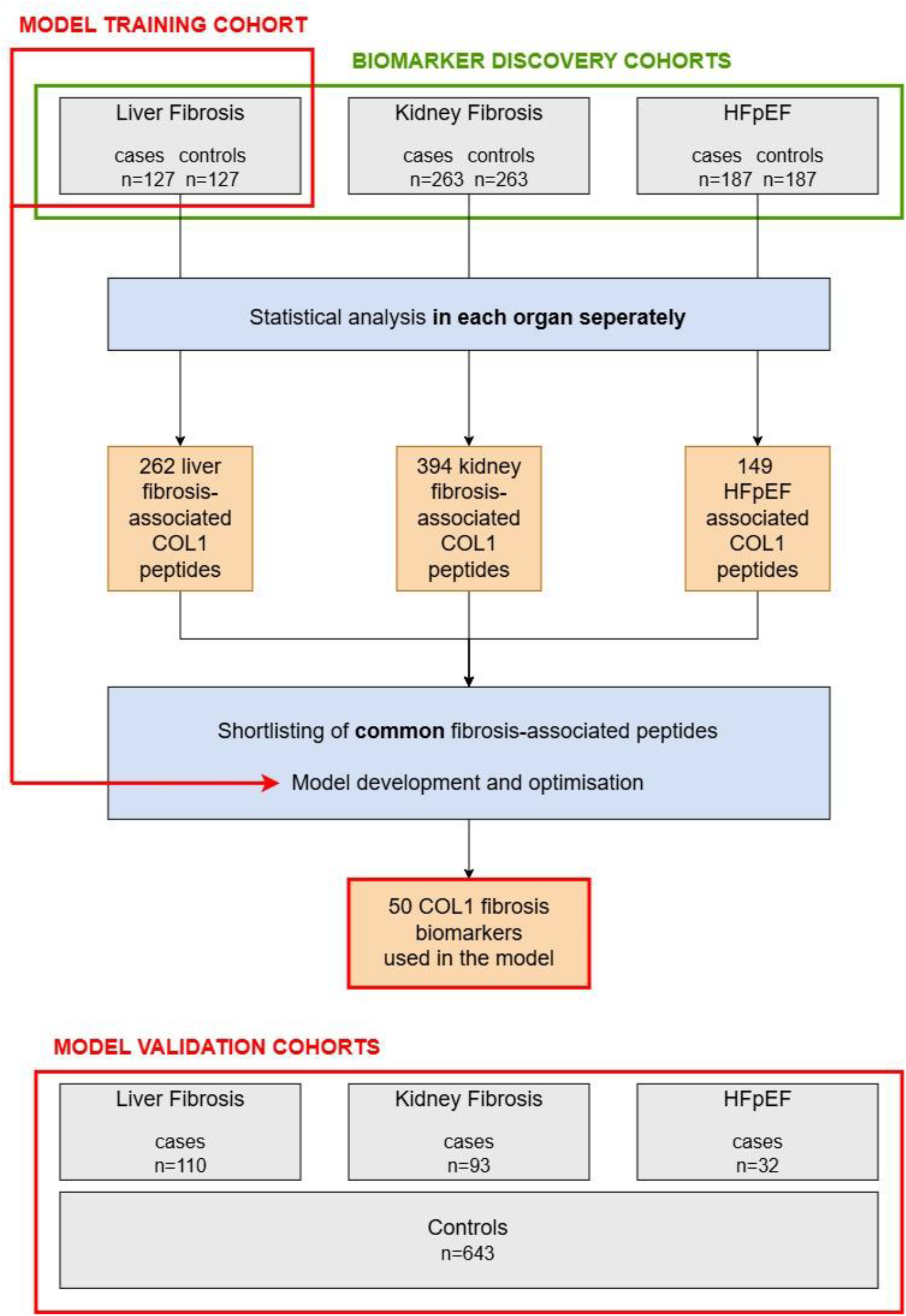
Study design. To identify fibrosis-associated COL1 peptides, separate case–control analyses were performed for liver fibrosis, kidney fibrosis, and heart failure with preserved ejection fraction (HFpEF). Peptides detected in ≥30% of samples in cases and/or controls were selected for each organ. Case–control comparisons were conducted using two approaches: without imputation of missing values and with missing values replaced by zero. Peptides were retained as organ-specific biomarkers if they showed a significant adjusted p-value in the analysis with imputation and a concordant fold change (FC) direction in the analysis without imputation. Peptides meeting the 30% frequency threshold across all three organs were considered candidate common fibrosis biomarkers and were further selected based on (i) statistical significance in at least two organ comparisons and (ii) consistent FC direction across all three. The top 60 peptides, ranked by Fisher’s combined p-values across organs, were further reduced to a final 50-peptide model using take-one-out cross-validation in the liver fibrosis cohort for biomarker discovery. A support vector machine (SVM) model was then trained based on these 50 fibrosis-associated peptides in the liver fibrosis cohort used for biomarker discovery. The model was validated in independent liver fibrosis, kidney fibrosis, and HFpEF cohorts. Abbreviations: COL1: collagen type I, HFpEF: heart failure with preserved ejection fraction.

Differences in model scores among the chronic disease and general population control cohorts were assessed using the Kruskal–Wallis test followed by Conover post hoc comparisons. The association between model scores and liver fibrosis stage was assessed using the Mann– Whitney test, while the correlation of the model scores with IFTA% was evaluated using Spearman’s correlation. These analyses were conducted using the kruskal.test, kwAllPairsConoverTest, wilcox.test and cor.test functions in R.

Data visualizations, including plots and heatmaps, were generated using the ggplot2 and gplots packages. R version 4.3.3 was used for all analyses and visualisations.

## Results

### Cohort characteristics

The case cohorts used for biomarker discovery are based on previously published studies, and were comprised of 127 patients with LDs (mainly cirrhosis), 263 patients with CKD (different aetiologies) and 187 patients with HF (HFpEF), along with the same number of matched control individuals. Detailed characteristics of these subjects are presented in **Table 1**. Of the 127 patients with LDs, 100 (78.6%) had fibrosis stage 4. The CKD cohort had a median IFTA of 30%, with an interquartile range (IQR) of 20-45%. Among the 187 datasets from patients with HF, 41 (21.9%) were classified as New York Heart Association (NYHA) class 1, 105 (56.1%) as NYHA class 2, 37 (19.8%) as NYHA class 3, and 4 (2.1%) as NYHA class 4. The same liver fibrosis cases with their matched controls were also used for the model training (**Figure 1**).

The model validation cohort was comprised of remaining subjects that could not be matched with controls during discovery and additional datasets generated after the publications of the initial studies. It included in total 235 cases and 643 controls. The cases were comprised of 110 patients with LDs (mainly cirrhosis, fibrosis stage 4), 93 patients with CKD (different aetiologies, median IFTA of 30%, [IQR 22.5-50%]) and 32 patients with HF (HFpEF, mainly NYHA stage 2 and 3). Detailed characteristics of the subjects are presented in **Table 1**.

### Liver fibrosis, kidney fibrosis and HFpEF share a common COL1 degradation signature

To date, 1097 naturally occurring COL1-derived peptides have been identified in urine, with 788 of them originating from COL1A1 and 309 from COL1A2. The 98.7% of these peptides are derived from the mature COL1A1 and COL1A2 chains, consequently reflecting the degradation of COL1. The abundance of these peptides was assessed in multiple cohorts, as shown in the study design in **Figure 1**.

Statistical analysis of cases and matched controls in the discovery cohorts identified 262 COL1 peptides associated with liver fibrosis (**Additional File 1; Supplementary Table 1**), 394 associated with kidney fibrosis (**Additional File 1; Supplementary Table 2**) and 149 associated with HFpEF (**Additional File 1; Supplementary Table 3**). Since many of these peptides were associated with more than one disease (**Additional File 2; Supplementary Figure 1**) the analysis resulted in 526 unique peptides, of which 403 were detected with a frequency greater than 30% (in either cases or controls) across all comparisons. Given differences in cohort sizes, and consequently statistical power, additional selection criteria were applied to ensure robustness. A subset of 90 peptides was identified based on the following conditions: (i) statistical significance in at least two comparisons and (ii) consistent fold change direction across all three comparisons. From this subset, the 60 peptides with the lowest Fisher’s combined p-values across organ comparisons were retained and subsequently reduced to a final set of 50 peptides using a take-one-out cross-validation approach (**Figure 2, Additional File 1; Supplementary Table 4**). The majority (n=45) peptides were derived from the COL1A1 chain, of which 33 were downregulated and 12 upregulated in fibrosis. The remaining 5 peptides were derived from COL1A2, with 3 being downregulated and 2 being upregulated in fibrosis (**Figure 2, Additional File 1; Supplementary Table 4**). All of the 50 fibrosis-associated peptides were fragments of the mature COL1A1 and COL1A2 chains, thus reflecting COL1 degradation.

**Figure 2.**
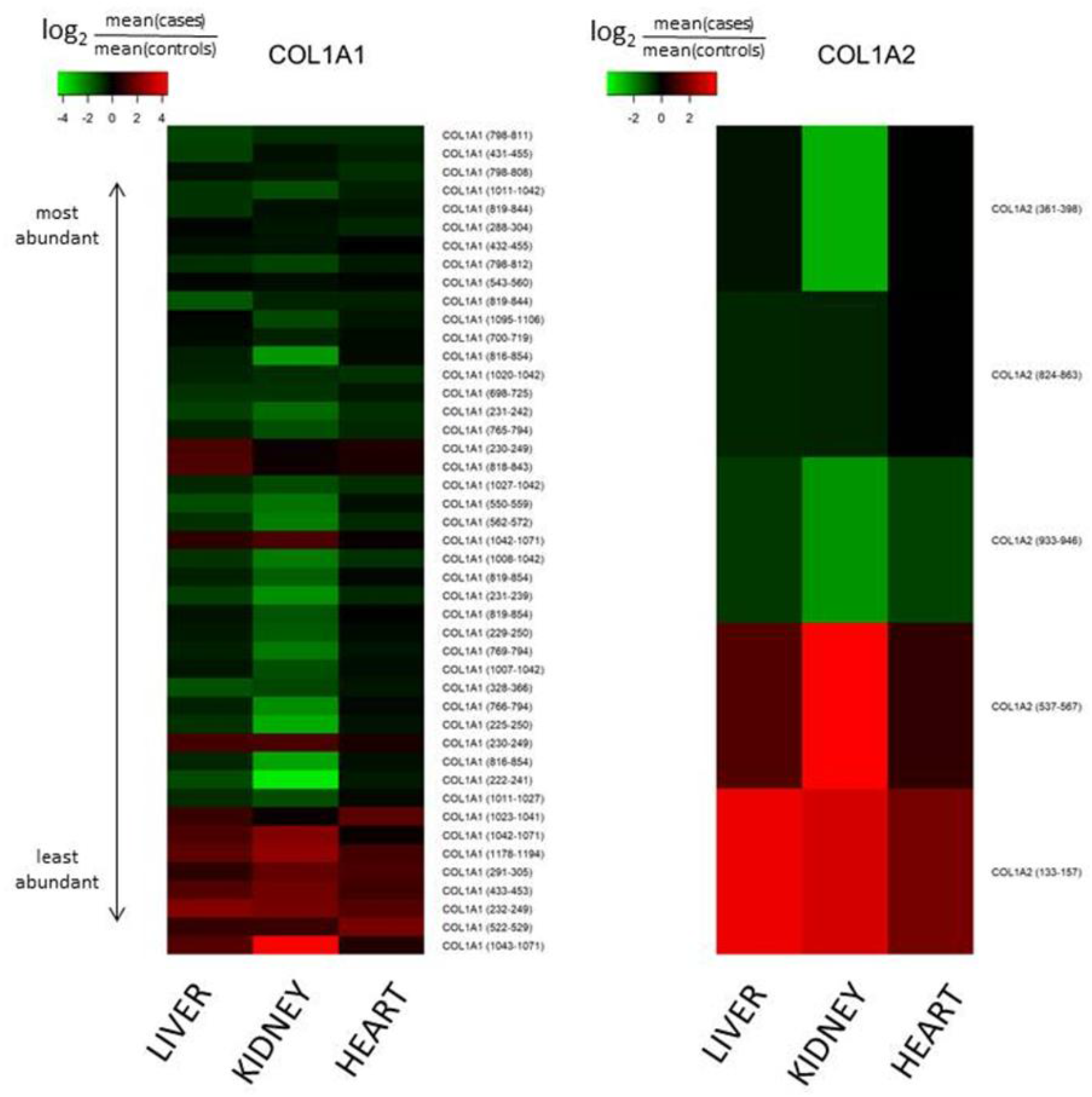
Regulation of urinary peptides in fibrosis. Heatmaps depicting the log2 fold change of the 50 fibrosis-associated urinary peptides across liver fibrosis, kidney fibrosis and heart failure with preserved ejection fraction (HFpEF) biomarker discovery cohorts. More than one peptide may share the same start and stop amino acid but differ in mass due to hydroxylation of proline or oxidation of methionine residues. The exact sequences of the fibrosis-associated peptides, including residue modifications are provided in **Additional File 1; Supplementary Table 4**. Peptides are ordered based on the median of their mean abundances in controls across the three organ comparisons, with the most abundant peptides displayed at the top of the heatmaps. Abbreviations: COL1A1: collagen type I alpha 1 chain, COL1A2: collagen type I alpha 2 chain.

### A model based on the common COL1 degradation signature is able to detect fibrosis, across different organs and diseases

An SVM model was generated using the final set of 50 COL1 degradation biomarkers identified through take-one-out cross-validation, with the liver fibrosis discovery cohort used as the training set (**Additional File 2; Supplementary Figure 2**). The model demonstrated good discriminatory performance, achieving an AUC of 0.935 (95% CI 0.917-0.953, p<0.0001) in the validation cohort for discriminating patients with all three chronic diseases from controls (**Figure 3A**). Importantly, the model maintained consistently high performance when evaluated separately in patients with liver fibrosis (n=110), kidney fibrosis (n=93), and HFpEF (n=32), with corresponding AUCs of 0.917 (95% CI 0.890-0.944, p<0.0001), 0.951 (95% CI 0.931-0.971, p<0.0001), and 0.950 (95% CI 0.903-0.997, p<0.0001), respectively, each compared against a shared control group (n=643) (**Figure 3B-D**). Comparison of these AUCs showed that discriminatory performance was significantly higher for kidney than liver fibrosis (p=0.026), while the differences between liver fibrosis and HFpEF and between kidney fibrosis and HFpEF were not significant (**Supplementary Table 5**).

**Figure 3.**
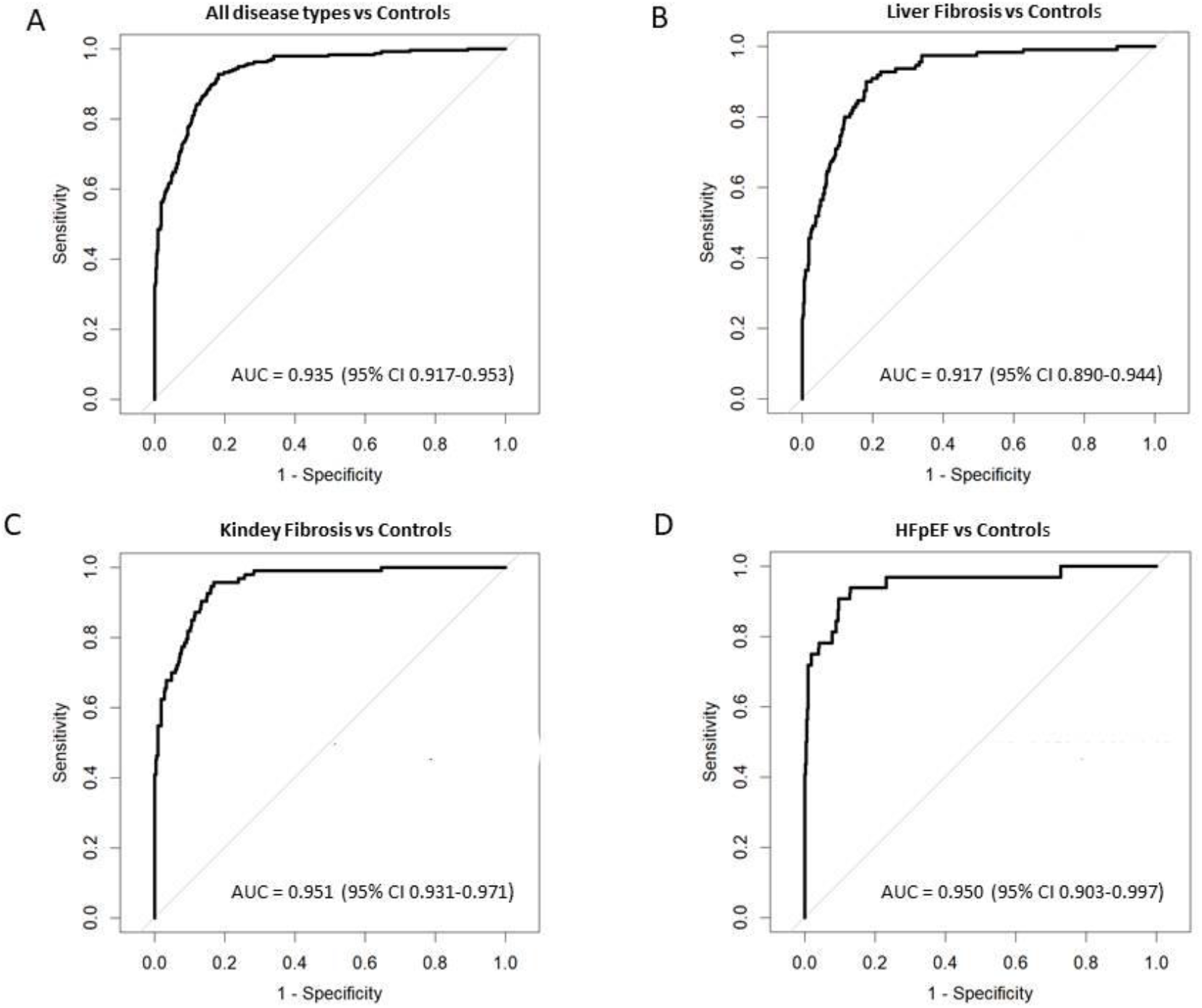
Performance of the collagen type I-based fibrosis model. (A) Receiver operating characteristic (ROC) curve showing model performance in the full validation cohort comprising patients with liver fibrosis, kidney fibrosis and HFpEF (n=235) and individuals from the general population (n=643). (B-D) ROC curves for organ-specific validation subgroups: liver fibrosis (n=110), kidney fibrosis (n=93), and HFpEF(n=32), each evaluated against a shared control group (n=643). Abbreviations: AUC: area under the receiver operating characteristic curve, CI: confidence interval, HFpEF: heart failure with preserved ejection fraction.

To assess whether the higher number of available controls compared to cases in the validation cohort influenced model performance, the analysis was repeated 100 times using balanced case-control datasets generated by random sampling, both for all disease groups combined and for each disease group separately. The mean AUCs were closely corresponding to those obtained using the full validation cohort (**Additional File 1; Supplementary Table 6**).

### Model scores based on the common COL1 degradation signature reflect fibrosis severity across organs

Consistent with the observed discriminatory performance, the distribution of model scores across fibrosis cohorts and general population controls is presented in **Figure 4A**. Model scores did not differ significantly among the liver fibrosis, kidney fibrosis, and HFpEF cohorts (Conover post hoc test, all p>0.05). In contrast, each disease cohort differed significantly from both general population control cohorts (all p<0.0001). Complete comparison results are provided in **Additional File 1; Supplementary Table 7.**

**Figure 4.**
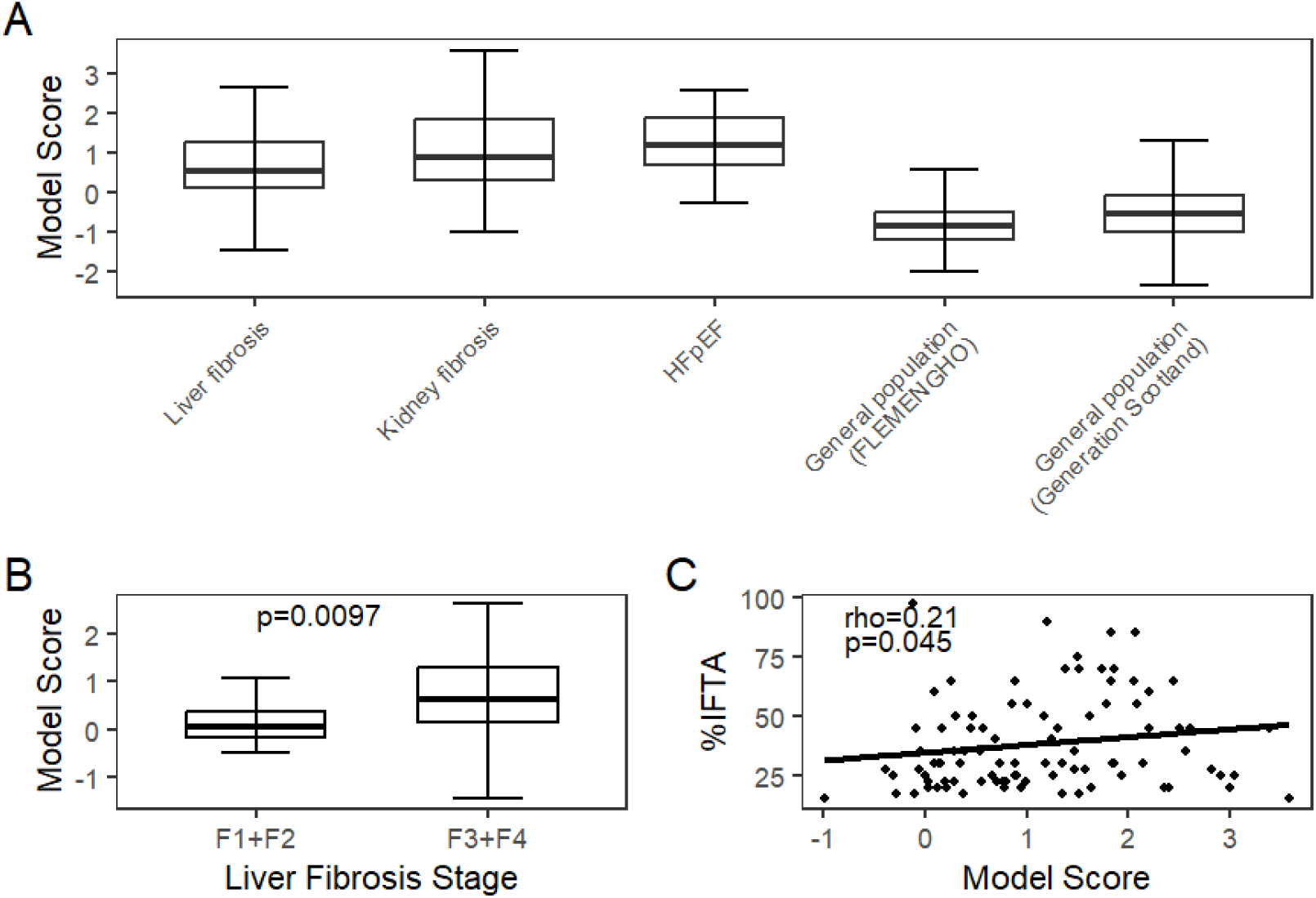
Distribution and clinical relevance of collagen type I-based fibrosis model scores. (A) Boxplots showing model scores across the validation cohorts: liver fibrosis (n=110), kidney fibrosis (n=93), heart failure with preserved ejection fraction (HFpEF, n=32), and two general population cohorts: FLEMENGHO (n=399) and Generation Scotland (n=244). Differences among the five groups were assessed using the Kruskal–Wallis test followed by Conover post hoc test. Scores did not differ significantly among the three disease groups (all Conover p > 0.05), while each fibrosis cohort differed significantly from both general population control cohorts (all Conover p<0.0001). Complete comparison results are provided in **Additional File 1; Supplementary Table 7**. (B) Boxplots of model scores stratified by fibrosis stage in the liver fibrosis validation cohort, comparing patients with stage F1-F2 (n=13) and stage F3–F4 (n=97) fibrosis. Group differences were evaluated using the Mann–Whitney test. (C) Scatter plot showing the correlation between model scores and the percentage of interstitial fibrosis and tubular atrophy (IFTA) in the kidney fibrosis validation cohort (n=93). Correlation was assessed using the Spearman’s test. Abbreviations: HFpEF: heart failure with preserved ejection fraction, IFTA: interstitial fibrosis and tubular atrophy, rho: Spearman’s correlation coefficient.

Furthermore, the model scores demonstrated a significant association with the liver and kidney fibrosis stage. Specifically, patients with advanced liver fibrosis (stage 3 or 4; n=97) had higher median scores of 0.62 [IQR 0.16 - 1.29], compared to those with early-stage fibrosis (stage 1 or 2; n=13), who had lower median scores of 0.05 [IQR -0.15 - 0.38] (p=0.0097; Mann–Whitney test; **Figure 4B**). Consistent with these findings, the model scores showed a significant positive correlation with the percentage of IFTA in the kidney fibrosis validation cohort (n=93; Spearman’s rho=0.21, p=0.045; **Figure 4C**).

## Discussion

Although increased COL1 synthesis is widely recognised as a major contributor to fibrosis^4–6^, the role of impaired collagen degradation has received much less attention, particularly when considering similarities across different chronic diseases. Analysis of the urinary peptidome provides a way to explore this aspect, as a large proportion of naturally occurring urinary peptides originating from COL1 is derived from the mature collagen rather than from the N- or C-terminal propeptides released during collagen synthesis^17^, indicating that they are products of COL1 degradation. Associations between urinary COL1 peptides and fibrotic disease in the liver^16^, kidney^17–19^ and heart^20,21^ have been reported previously, supporting their relevance in this context. On this basis, COL1-derived peptides have been also used to develop models for the non-invasive assessment of fibrosis in the liver^16^ and kidney^22^ (comparison with the common fibrosis model in **Additional File 1; Supplementary Table 8**). Moreover, our previous study^19^, focusing on COL1A1-derived (*i.e.* the specific COL1A1 chain) urinary peptides, showed that early stage fragments in the collagen degradation process, likely directly derived from the COL1A1 molecule as a result of endopeptidase activity, were consistently downregulated in patients with CKD, suggesting that the accumulation of COL1 in fibrosis is partly the result of an attenuated COL1 degradation process^19^. However, a small proportion of peptides from later degradation steps, likely derived from larger peptides through exopeptidase activity, were upregulated^19^, which might explain why the majority of the 50 common COL1-derived biomarkers identified in the present study were downregulated in fibrosis, while a smaller proportion were upregulated. A study using a unilateral ureteral obstruction mouse model further supported the hypothesis of a reduced COL1 degradation during fibrosis development, showing that COL1-degradation products collected from the pelvis of the obstructed (fibrotic) kidney were predominantly downregulated compared with non-fibrotic controls^44^. Importantly, this downregulation was already observed three days after obstruction, when an increase in collagen protein deposition in the kidney tissue was not yet detectable^44^. These findings suggest that impaired collagen degradation may contribute to the early development of kidney fibrosis rather than occurring solely as a consequence of established fibrosis^44^. In the current study we investigated the association of COL1 peptides with each fibrotic disease (LDs, CKD and HF) separately and identified peptides that were commonly associated with all three diseases. All of these peptides were fragments of the mature COL1, therefore products of COL1 degradation, and most of them were downregulated in fibrosis, further supporting the concept of reduced collagen degradation during fibrotic remodelling across different tissues.

A plausible explanation for reduced collagen cleavage is the increased cross-linking of collagen fibres in fibrotic tissue. This may be driven by the upregulation of cross-linking enzymes, including lysyl hydroxylase^45,46^, lysyl oxidase^47^ and transglutaminase^48^, and the accumulation of advanced glycation end-products^49–51^ in fibrotic disease. These alterations increase collagen stability and limit its accessibility to MMPs, thereby promoting extracellular matrix accumulation^13,45,48,51^. Moreover, given the dependence of MMP catalytic activity on zinc, collagen degradation may be further hindered by the consistent downregulation of metallothioneins, resulting in reduced zinc ion availability across fibrotic diseases^52^.

Chronic diseases, including LDs^53^, CKD^54^ and HF^55^ are major contributors of global morbidity and mortality. However, commonly used clinical tests that assess liver function (LFTs), kidney function (eGFR), and heart injury (troponins or natriuretic peptides) often detect these diseases at late stages, when significant organ damage has occurred, with limited means to stop the progression^56–60^. Therefore, there is a clear clinical need for tools that would enable the early detection of chronic diseases, when lifestyle and pharmacological interventions are more effective at slowing disease progression. Given that fibrosis develops from the early stages of these conditions and progressively contributes to organ dysfunction, its detection may offer an opportunity for earlier therapeutic intervention across chronic diseases^59,61,62^.

Currently the gold standard for the diagnosis of fibrosis is biopsy, an invasive procedure that in some cases leads to health complications and is prone to sampling bias. Therefore, there is a need for the development of methods for the non-invasive assessment of fibrosis. Various imaging and blood/urine-based methods have been developed towards this end. Among imaging techniques, vibration-controlled transient elastography (FibroScan) is the most validated method for liver fibrosis assessment, although confounding factors must be considered to avoid misinterpretation of fibrosis severity^24,63^. For the assessment of cardiac fibrosis, late gadolinium enhancement cardiac magnetic resonance is used in clinical practice, however, it has limited sensitivity for detecting diffuse interstitial fibrosis and requires the administration of gadolinium-based contrast agents^27,64^. Although imaging-based methods such as ultrasound, positron emission tomography/computed tomography, and magnetic resonance imaging show potential for the non-invasive assessment of renal fibrosis, their use in clinical practice remains limited^65^. Blood-based tests such as aspartate aminotransferase to platelet ratio index and fibrosis-4 index are widely used for liver fibrosis but represent indirect markers influenced by confounders^24,63^. Similarly, circulating biomarkers such as galectin-3 and soluble ST2 have been associated with cardiac fibrosis, yet lack specificity due to elevation in other inflammatory conditions^27^. Proposed biomarkers for kidney fibrosis, such as human epididymis protein 4 (HE4) and dickkopf-related protein 3 (DKK3), also reflect broader biological processes beyond fibrosis^59^. In contrast, biomarkers derived from collagen turnover may provide a more direct assessment of fibrosis. Previous studies have shown that collagen derived peptides not only provide mechanistic insight into the fibrotic process but also could help in the detection and prediction of fibrotic disease. For example, the enhanced liver fibrosis (ELF) test is a widely validated blood test based on the procollagen type III N-terminal propeptide (PIIINP), reflecting collagen type III synthesis, hyaluronic acid and tissue inhibitor of metalloproteinase 1 ^63^. A meta-analysis demonstrated good diagnostic performance of the ELF test in both significant fibrosis (≥F2) and advanced fibrosis (≥F3), with summary receiver operating characteristic AUCs of 0.81 and 0.83, respectively^66^. The levels of PIIINP in urine are also correlated with the degree of kidney fibrosis^59^. Additionally, the procollagen type I C-terminal propeptide (PICP), reflecting synthesis of COL1, is associated with myocardial fibrosis and can predict adverse outcome in HF patients, while, the cross-linked collagen type I C-terminal telopeptide (CITP), released by the collagenase matrix metalloproteinase-1 and reflecting COL1 degradation, predicts HFpEF incidence^67^. In addition, urinary peptide models including also collagen fragments have demonstrated ability to detect fibrosis in the liver^16^ and kidney^22^.

Building on these observations, the present study identified a common urinary COL1 degradation signature across patients with fibrosis affecting different organs using peptidomics. A model developed based on this signature was able to assess fibrosis in an independent validation cohort, independently of its specific organ localisation. These findings support the concept that fibrosis is driven by shared extracellular matrix remodelling processes across different tissues. Since fibrosis could be interpreted as disturbance in COL1 homeostasis resulting from both an increased COL1 production and a decreased COL1 degradation, a model based solely on degradation products is unlikely to fully capture the complexity of the disease. Therefore, using a combination of the COL1 degradation-based model with markers of COL1 synthesis may be the most beneficial strategy for fibrosis detection.

The study has several limitations. First, because no urine samples from patients with confirmed heart fibrosis were available, samples from patients with HFpEF were investigated based on the assumption that HFpEF is tightly linked to cardiac fibrosis. This approach is supported by a previous study by Hahn et al.^38^, who analysed myocardial tissue from a large cohort of HFpEF patients and found that 93% exhibited cardiac fibrosis. Moreover, because most LDs and CKD patients used in the study had already late-stage disease, future validation of the model will be needed to determine its performance in detecting fibrosis at early stages. The low eGFR in the CKD discovery cohort cannot be matched with the controls, as low eGFR defines CKD. Influence of impaired kidney function on the urinary peptide levels in this cohort cannot be formally excluded, however peptide intensities are normalized using 29 internal standard collagen fragments that are generally unaffected by disease. As such it is reasonable to assume that reduced eGFR does not affect the normalised signal intensities.

Controls from population-based studies with confirmed absence of fibrosis were also not available, since in clinical practice fibrosis is assessed based on specific clinical indications. Therefore, the possibility that some control individuals had underlying fibrosis despite the absence of clinical symptoms, resulting in misclassification and affecting the classifier’s specificity, cannot be excluded. However, their clinical characteristics suggest that the majority were unlikely to have significant fibrotic disease, supporting their use as comparator groups in the analyses. Additionally, the exact mechanisms underlying the generation of COL1-derived urinary biomarkers are not fully understood. Although in a previous study we modelled the generation of COL1 peptides as the result of protease activity^19^, the precise tissue origin of the peptides cannot be traced. Accordingly, these biomarkers reflect systemic collagen turnover, which may also be influenced by processes beyond fibrosis. Nevertheless, a previous study identifying liver fibrosis biomarkers showed gradual changes in the abundance of urinary collagen-derived peptides with liver disease severity, suggesting that the observed changes reflect fibrosis in liver^16^. Moreover, the systemic nature of the common fibrosis biomarkers identified in our study may represent an advantage for fibrosis detection, as fibrosis can be identified independently of the organ of origin. In addition, the cross-sectional design and absence of longitudinal follow-up data limit conclusions regarding the causal relationship between altered COL1 degradation and fibrosis development and progression. Another limitation is that comparison with other fibrosis biomarkers could not be performed, as these biomarkers were not measured in the investigated cohorts.

Future studies should validate the model in patients with early-stage fibrosis and in at-risk populations, including individuals with ageing-related risk factors, hypertension, obesity, diabetes and chronic low-grade inflammation. These studies should also compare its performance with established fibrosis markers and investigate whether combining the model with existing markers, particularly markers of collagen synthesis, further improves its performance. Ultimately, prospective clinical trials in at-risk populations could investigate whether individuals with a positive fibrosis signature before clinically apparent organ damage benefit from earlier preventive or therapeutic interventions with potential anti-fibrotic effects, such as sodium-glucose cotransporter 2 (SGLT2) inhibitors, or from closer clinical monitoring. In addition, such a signature could potentially facilitate the development and evaluation of anti-fibrotic therapies by enabling biomarker-based patient stratification according to shared molecular mechanisms rather than clinical symptoms, similar to approaches used in oncology^68^.

## Conclusion

By investigating naturally occurring peptides in urine, the study revealed a common COL1 degradation signature across different diseases with fibrotic component, including LDs, CKD and HF. A model based on this signature could detect fibrosis in an independent validation cohort, highlighting the potential of urinary biomarkers as a tool for a non-invasive assessment of fibrosis, irrespective of organ origin. The predominance of downregulated COL1 degradation products in this signature further supports the hypothesis that attenuated COL1 degradation contributes to fibrotic remodelling across different organs. Rather than replacing organ-specific diagnostics, this approach could contribute to the early identification of fibrotic remodelling and support treatment selection before irreversible organ damage occurs.

## Data Availability

The datasets used and/or analysed during the current study are available from the corresponding author on reasonable request.

## List of abbreviations

AUC: Area under the receiver operating characteristic curve
CE-MS: Capillary electrophoresis coupled to mass spectrometry
CI: Confidence interval
CITP: Collagen type I C-terminal telopeptide
CKD: Chronic kidney disease
COL1: Collagen type I
COL1A1: Collagen type I alpha 1 chain
COL1A2: Collagen type I alpha 2 chain
DKK3: Dickkopf-related protein 3
ELF: Enhanced liver fibrosis
eGFR: Estimated glomerular filtration rate
HE4: Human epididymis protein 4
HF: Heart failure
HFpEF: Heart failure with preserved ejection fraction
IFTA: Interstitial fibrosis and tubular atrophy
IQR: Interquartile range
LDs: Liver diseases
LFTs: Liver function tests
MMP: Matrix metalloproteinase
NYHA: New York Heart Association
PICP: Procollagen type I C-terminal propeptide
PIIINP: Procollagen type III N-terminal propeptide
SGLT2: Sodium-glucose cotransporter 2
SVM: Support vector machine

## Declarations

### Ethics approval and consent to participate

The Study was conducted according to the guidelines of the Declaration of Helsinki. For the CKD cohorts the local ethics committee of the Friedrich-Alexander Universitaet Erlangen-Nuernberg provided approval for the nephrological biobank of the Klinikum Bayreuth (ethic approval code 264_20 B) and the urinary proteomics analysis (ethic approval code 221_20 B). For the liver cohort (TENDENCY), the study was approved by Northeast-York research ethics committee, REC reference 19/NE/0213, IRAS project ID 260179. For the cohort from Bannaga et al.: the study was approved by Coventry and Warwickshire and North East-York Research National Health Service Ethics Committees UK (Reference numbers 09/H1211/38 and 19/NE/0213), and Ethics Committee of the Medical School Hannover (Reference number: 901). For the HFpEF cohort approval was provided by the ethics committee of the University Aachen (EK163/19). In addition, ethical review and approval were not required for this study due to all data being fully anonymized, based on the opinion of the ethics committee of the Hannover Medical School, Germany (no. 3116-2016).

## Consent for publication

Not applicable.

## Author contributions

IKM contributed to the study design and conceptualisation, performed most of the data analysis, contributed to data interpretation and wrote the manuscript. YH, LC, HR, JB, JAS, JM, FP, PR, CD, and AB contributed to the acquisition and interpretation of the data. JS contributed to part of the data analysis. AV contributed to the study design and conceptualisation. RPA contributed to the study design and conceptualisation and to the acquisition and interpretation of the data. JPS, HM, and AL contributed to the study design and conceptualisation and performed substantial revision of the manuscript. All authors read and approved the final manuscript.

## Acknowledgements

Not applicable

## Additional Material

**Additional File 1**(.xls) contains Supplementary Tables 1-8.

**Supplementary Table 1:** 262 liver fibrosis associated COL1 peptides.

**Supplementary Table 2:** 394 kidney fibrosis associated COL1 peptides.

**Supplementary Table 3:** 149 heart failure with preserved ejection fraction (HFpEF) associated COL1 peptides.

**Supplementary Table 4:** 50 COL1 fibrosis biomarkers.

**Supplementary Table 5:** Differences in AUC were assessed for each combination of organ-specific validation cohorts (liver fibrosis vs. kidney fibrosis, liver fibrosis vs. heart failure with preserved ejection fraction (HFpEF), and kidney fibrosis vs. HFpEF) using nonparametric bootstrap resampling with 10000 replicates. For each replicate, cases from the two organ groups being compared were sampled independently with replacement, while the shared controls were sampled once with replacement and used for both ROC curves, thus preserving the shared control structure of the original analysis. The original numbers of cases and controls were maintained in each replicate. For each bootstrap replicate, the difference between the two resulting AUCs was calculated, and the standard deviation of the 10000 bootstrap AUC differences was determined. Following the bootstrap testing approach implemented in the pROC package, the AUC difference observed in the original data was divided by the standard deviation of the bootstrap AUC differences to obtain the test statistic (D), which was compared with the standard normal distribution to obtain a two-sided p-value.

**Supplementary Table 6:** Model performance using balanced case-control datasets. Controls were randomly selected to match the number of cases, followed by assessment of model performance. Control selection and subsequent analysis were repeated 100 times for each comparison (all disease types combined, liver fibrosis, kidney fibrosis and heart failure with preserved ejection fraction (HFpEF)). Mean, minimum and maximum AUC and specificity across the 100 repetitions are presented. Sensitivity and specificity were determined using the predefined model cut-off of >0.045. Sensitivity remained unchanged across repetitions, as all cases were retained and only controls were randomly selected.

**Supplementary Table 7:** Comparison of fibrosis model scores among cohorts. Differences in model scores among the liver fibrosis, kidney fibrosis, heart failure with preserved ejection fraction (HFpEF), and two general population control cohorts (FLEMENGHO and Generation Scotland) were assessed using the Kruskal–Wallis test (p-value < 2.2E-16), followed by Conover post hoc comparisons.

**Supplementary Table 8:** Sensitivity and specificity analysis based on samples from the validation cohort. For the existing models, previously established cut-offs (BH29^22^ > 0.025, LivFib50^16^ > -0.02) were applied, while for the general fibrosis model, the cut-off was determined using the Youden index and set to > 0.045 based on the total cross-validated scores in the training cohort.

Small letters in peptide sequences symbolise post-translational modifications (p: hydroxyproline, k: hydroxylysine, m: oxidized methionine)

**Additional File 2**(.docx) contains Supplementary Figures 1-2.

**Supplementary Figure 1:** Venn diagram showing the overlap of fibrosis-associated COL1 peptides across different organ case-control comparisons.

**Supplementary Figure 2.** Performance of the collagen type I-based fibrosis model during cross-validation. (A) Receiver operating characteristic (ROC) curve showing the performance of the 50-peptide model based on the cross-validation scores in the liver fibrosis training cohort, comprising patients with liver fibrosis (n=127) and matched controls (n=127). The model achieved an AUC of 0.878 (95% CI 0.831-0.915, p<0.0001). The optimal cut-off of >0.045 was determined using the Youden index and resulted in a sensitivity of 77.2% (95% CI 68.9-84.1%) and specificity of 89.0% (95% CI 82.2-93.8%). (B) Box plots showing the cross-validation model scores in patients with liver fibrosis and matched controls. Model scores were significantly higher in patients with liver fibrosis compared with controls (p<0.0001, Mann–Whitney test). Abbreviations: AUC: area under the receiver operating characteristic curve, CI: confidence interval.

## References

1 Lee CJM, Kosyakovsky LB, Khan MS, Wu F, Chen G, Hill JA et al. Cardiovascular, Kidney, Liver, and Metabolic Interactions in Heart Failure: Breaking Down Silos. Circ Res 2025; 136: 1170–1207.

2 Wynn TA, Ramalingam TR. Mechanisms of fibrosis: Therapeutic translation for fibrotic disease. Nat Med 2012; 18: 1028–1040.

3 Zhao X, Kwan JYY, Yip K, Liu PP, Liu FF. Targeting metabolic dysregulation for fibrosis therapy. Nat Rev Drug Discov 2020; 19: 57–75.

4 Thompson KJ, Mckillop IH, Schrum LW. Targeting collagen expression in alcoholic liver disease. World J Gastroenterol 2011; 17: 2473–2481.

5 Bülow RD, Boor P. Extracellular Matrix in Kidney Fibrosis: More Than Just a Scaffold. J Histochem Cytochem 2019; 67: 643–661.

6 Parichatikanond W, Luangmonkong T, Mangmool S, Kurose H. Therapeutic targets for the treatment of cardiac fibrosis and cancer: Focusing on tgf-β Signaling. Front Cardiovasc Med 2020; 7: 1–19.

7 Antar SA, Ashour NA, Marawan ME, Al-Karmalawy AA. Fibrosis: Types, Effects, Markers, Mechanisms for Disease Progression, and Its Relation with Oxidative Stress, Immunity, and Inflammation. Int J Mol Sci 2023; 24: 4004.

8 Kim KK, Sheppard D, Chapman HA. TGF-β1 signaling and tissue fibrosis. Cold Spring Harb Perspect Biol 2018; 10: 1–34.

9 McKleroy W, Lee TH, Atabai K. Always cleave up your mess: Targeting collagen degradation to treat tissue fibrosis. Am J Physiol - Lung Cell Mol Physiol 2013; 304: L709–L721.

10 Devos H, Zoidakis J, Roubelakis MG, Latosinska A, Vlahou A. Reviewing the Regulators of COL1A1. Int J Mol Sci 2023; 24: 7480.

11 Shan L, Wang F, Zhai D, Meng X, Liu J, Lv X. Matrix metalloproteinases induce extracellular matrix degradation through various pathways to alleviate hepatic fibrosis. Biomed Pharmacother 2023; 161: 114472.

12 Verzijl N, DeGroot J, Thorpe SR, Bank RA, Shaw JN, Lyons TJ et al. Effect of collagen turnover on the accumulation of advanced glycation end products. J Biol Chem 2000; 275: 39027–39031.

13 Panwar P, Butler GS, Jamroz A, Azizi P, Overall CM, Brömme D. Aging-associated modifications of collagen affect its degradation by matrix metalloproteinases. Matrix Biol 2018; 65: 30–44.

14 Makareeva E, Leikin S. Collagen Structure, Folding and Function. Shapiro JR Byers PH Glorieux FH Sponselle PD Ed Osteogenes Imperfecta Acad Press 2014: 71–84.

15 Mavrogeorgis E, Mischak H, Latosinska A, Siwy J, Jankowski V, Jankowski J. Reproducibility evaluation of urinary peptide detection using CE-MS. Molecules 2021; 26: 7260.

16 Bannaga AS, Metzger J, Kyrou I, Voigtländer T, Book T, Melgarejo J et al. Discovery, validation and sequencing of urinary peptides for diagnosis of liver fibrosis—A multicentre study. EBioMedicine 2020; 62: 103083.

17 Mavrogeorgis E, Mischak H, Latosinska A, Vlahou A, Schanstra JP, Siwy J et al. Collagen-derived peptides in CKD: A link to fibrosis. Toxins 2022; 14: 1–13.

18 Good DM, Zürbig P, Argilés À, Bauer HW, Behrens G, Coon JJ et al. Naturally occurring human urinary peptides for use in diagnosis of chronic kidney disease. Mol Cell Proteomics 2010; 9: 2424–2437.

19 Mina IK, Iglesias-Martinez LF, Ley M, Fillinger L, Perco P, Siwy J et al. Investigation of the Urinary Peptidome to Unravel Collagen Degradation in Health and Kidney Disease. Proteomics 2025; 25: e202400279.

20 He T, Mischak M, Clark AL, Campbell RT, Delles C, Díez J et al. Urinary peptides in heart failure: a link to molecular pathophysiology. Eur J Heart Fail 2021; 23: 1875–1887.

21 He T, Melgarejo JD, Clark AL, Yu Y, Thijs L, Díez J et al. Serum and urinary biomarkers of collagen type-I turnover predict prognosis in patients with heart failure. Clin Transl Med 2021; 11: 2–5.

22 Catanese L, Siwy J, Mavrogeorgis E, Amann K, Mischak H, Beige J et al. A novel urinary proteomics classifier for non-invasive evaluation of interstitial fibrosis and tubular atrophy in chronic kidney disease. Proteomes 2021; 9: 32.

23 Ku JC, Raiten J, Li Y. Understanding fibrosis: Mechanisms, clinical implications, current therapies, and prospects for future interventions. Biomed Eng Adv 2024; 7: 100118.

24 Canivet CM, Boursier J. Screening for Liver Fibrosis in the General Population: Where Do We Stand in 2022? Diagnostics 2023; 13: 1–15.

25 Abdelhameed F, Kite C, Lagojda L, Dallaway A, Chatha KK, Chaggar SS et al. Non-invasive Scores and Serum Biomarkers for Fatty Liver in the Era of Metabolic Dysfunction-associated Steatotic Liver Disease (MASLD): A Comprehensive Review From NAFLD to MAFLD and MASLD. Curr Obes Rep 2024; 13: 510–531.

26 Wan S, Wang S, He X, Song C, Wang J. Noninvasive diagnosis of interstitial fibrosis in chronic kidney disease: a systematic review and meta-analysis. Ren Fail 2024; 46: 2367021.

27 Zhu L, Wang Y, Zhao S, Lu M. Detection of myocardial fibrosis: Where we stand. Front Cardiovasc Med 2022; 9: 926378.

28 Mischak H, Vlahou A, Ioannidis JPA. Technical aspects and inter-laboratory variability in native peptide profiling: The CE-MS experience. Clin Biochem 2013; 46: 432–443.

29 Latosinska A, Siwy J, Mischak H, Frantzi M. Peptidomics and proteomics based on CE-MS as a robust tool in clinical application: The past, the present, and the future. Electrophoresis 2019; 40: 2294–2308.

30 Jantos-Siwy J, Schiffer E, Brand K, Schumann G, Rossing K, Delles C et al. Quantitative urinary proteome analysis for biomarker evaluation in chronic kidney disease. J Proteome Res 2009; 8: 268–281.

31 Martens DS, Thijs L, Latosinska A, Trenson S, Siwy J, Zhang ZY et al. Urinary peptidomic profiles to address age-related disabilities: a prospective population study. Lancet Healthy Longev 2021; 2: e690–e703.

32 Mina IK, Mavrogeorgis E, Siwy J, Stojanov R, Mischak H, Latosinska A et al. Multiple urinary peptides display distinct sex-specific distribution. Proteomics 2024; 24: e2300227.

33 Rossing K, Mischak H, Dakna M, Zu P, Novak J, Julian BA et al. Urinary Proteomics in Diabetes and CKD. J Am Soc Nephrol 2008;: 1283–1290.

34 Tofte N, Lindhardt M, Adamova K, Bakker SJL, Beige J, Beulens JWJ et al. Early detection of diabetic kidney disease by urinary proteomics and subsequent intervention with spironolactone to delay progression (PRIORITY): a prospective observational study and embedded randomised placebo-controlled trial. Lancet Diabetes Endocrinol 2020; 8: 301–312.

35 Zhang Z, Staessen JA, Thijs L, Gu Y, Liu Y, Jacobs L et al. Left ventricular diastolic function in relation to the urinary proteome: A proof-of-concept study in a general population. Int J Cardiol 2014; 176: 158–165.

36 Smith BH, Campbell A, Linksted P, Fitzpatrick B, Jackson C, Kerr SM et al. Cohort profile: Generation scotland: Scottish family health study (GS: SFHS). The study, its participants and their potential for genetic research on health and illness. Int J Epidemiol 2013; 42: 689–700.

37 Mohammed SF, Hussain S, Mirzoyev SA, Edwards WD, Maleszewski JJ, Redfield MM. Coronary microvascular rarefaction and myocardial fibrosis in heart failure with preserved ejection fraction. Circulation 2015; 131: 550–559.

38 Hahn VS, Yanek LR, Vaishnav J, Ying W, Vaidya D, Lee YZJ et al. Endomyocardial Biopsy Characterization of Heart Failure With Preserved Ejection Fraction and Prevalence of Cardiac Amyloidosis. JACC Heart Fail 2020; 8: 712–724.

39 Hussain Y, Bannaga A, Fisher N, Krishnamoorthy A, Kimani P, Malik A et al. The Fatty Liver, Cirrhosis, and Liver Cancer Study (TENDENCY): Protocol for a Multicenter Case-Control Study. JMIR Res Protoc 2023; 12: 1–7.

40 Frantzi M, Keller F, Latosinska A, Beige J, Mebazaa A, Caillard A et al. Urinary Collagen Peptides Predict Mortality. Proteomics 2026;: e70131.

41 Ho DE, Imai K, King G, Stuart EA. MatchIt: Nonparametric Preprocessing for Parametric Causal Inference. J Stat Softw 2011; 42: 1–28.

42 An DW, Yu YL, Martens DS, Latosinska A, Zhang ZY, Mischak H et al. Statistical approaches applicable in managing OMICS data: Urinary proteomics as exemplary case. Mass Spectrom Rev 2023;: 1–18.

43 Benjamini Y, Hochberg Y. Controlling the false discovery rate: a practical and powerful approach to multiple testing. J R Stat Soc Ser B Methodol 1995; 57: 289–300.

44 Frattini T, Breuil B, Buléon M, Feuillet G, Chabbert M, Delecroix E et al. Reduced collagen degradation in pelvic urine precedes kidney fibrosis induced by unilateral ureteral obstruction. doi:10.21203/rs.3.rs-6872444/v1.

45 Piersma B, Bank RA. Collagen cross-linking mediated by lysyl hydroxylase 2: An enzymatic battlefield to combat fibrosis. Essays Biochem 2019; 63: 377–387.

46 Remst DFG, Blaney Davidson EN, Vitters EL, Blom AB, Stoop R, Snabel JM et al. Osteoarthritis-related fibrosis is associated with both elevated pyridinoline cross-link formation and lysyl hydroxylase 2b expression. Osteoarthritis Cartilage 2013; 21: 157–164.

47 Zhang XQ, Li X, Zhou WQ, Liu X, Huang JL, Zhang YY et al. Serum Lysyl Oxidase Is a Potential Diagnostic Biomarker for Kidney Fibrosis. Am J Nephrol 2020; 51: 907–918.

48 Johnson TS, El-Koraie AF, Skill NJ, Baddour NM, El Nahas AM, Njloma M et al. Tissue transglutaminase and the progression of human renal scarring. J Am Soc Nephrol 2003; 14: 2052– 2062.

49 Koska J, Gerstein HC, Beisswenger PJ, Reaven PD. Advanced Glycation End Products Predict Loss of Renal Function and High-Risk Chronic Kidney Disease in Type 2 Diabetes. Diabetes Care 2022; 45: 684–691.

50 Makino H, Shikata K, Hironaka K, Kushiro M, Yamasaki Y, Sugimoto H et al. Ultrastructure of nonenzymatically glycated mesangial matrix in diabetic nephropathy. Kidney Int 1995; 48: 517– 526.

51 DeGroot J, Verzijl N, Budde M, Bijlsma JWJ, Lafeber FPJG, TeKoppele JM. Accumulation of advanced glycation end products decreases collagen turnover by bovine chondrocytes. Exp Cell Res 2001; 266: 303–310.

52 Campos MAJ, Stroggilos R, Schanstra J-P, Vlahou A, Brunet M, Jonas J-C et al. Pan-fibrotic gene expression signature in major chronic diseases by integrative bulk and single-cell transcriptomic analyses. medRxiv 2025;: 10.1101/2025.10.07.25337472.

53 Gan C, Yuan Y, Shen H, Gao J, Kong X, Che Z et al. Liver diseases: epidemiology, causes, trends and predictions. Signal Transduct Target Ther 2025; 10: 33.

54 Ying M, Shao X, Qin H, Yin P, Lin Y, Wu J et al. Disease Burden and Epidemiological Trends of Chronic Kidney Disease at the Global, Regional, National Levels from 1990 to 2019. Nephron 2023; 148: 113–123.

55 Shahim B, Kapelios CJ, Savarese G, Lund LH. Global Public Health Burden of Heart Failure: An Updated Review. Card Fail Rev 2023; 9: e11.

56 Ahmed Z, Ahmed U, Walayat S, Ren J, Martin DK, Moole H et al. Liver function tests in identifying patients with liver disease. Clin Exp Gastroenterol 2018; 11: 301–307.

57 Ginès P, Castera L, Lammert F, Graupera I, Serra-Burriel M, Allen AM et al. Population screening for liver fibrosis: Toward early diagnosis and intervention for chronic liver diseases. Hepatology 2022; 75: 219.

58 Rende U, Guller A, Goldys EM, Pollock C, Saad S. Diagnostic and prognostic biomarkers for tubulointerstitial fibrosis. J Physiol 2023; 601: 2801–2826.

59 Rupprecht H, Catanese L, Amann K, Hengel FE, Huber TB, Latosinska A et al. Assessment and Risk Prediction of Chronic Kidney Disease and Kidney Fibrosis Using Non-Invasive Biomarkers. Int J Mol Sci 2024; 25: 1–18.

60 Fang X, Xu Y, Zhao Y, Li L, Cheng Z, Zhang Z, et al. Beyond Traditional Screening: The Future of Heart Failure Detection With Biomarkers and AI. INew Med; n/a: e70051.

61 Pose E, Piano S, Thiele M, Fabrellas N, Tsochatzis EA, Ginès P. Moving diagnosis of liver fibrosis into the community. J Hepatol 2025; 83: 258–270.

62 Ureche C, Nedelcu A-E, Sascău RA, Stătescu C, Kanbay M, Covic A. Role of collagen turnover biomarkers in the noninvasive assessment of myocardial fibrosis: an update. Biomark Med 2020; 14: 1265–1275.

63 Castera L. Noninvasive Assessment of Liver Fibrosis. Dig Dis 2015; 33: 498–503.

64 Mewton N, Liu CY, Croisille P, Bluemke D, Lima JAC. Assessment of myocardial fibrosis with cardiovascular magnetic resonance. J Am Coll Cardiol 2011; 57: 891–903.

65 Yuan T, Wang H, Kang T, Wu W, Ou S. Advancements in the non-invasive diagnosis of renal fibrosis. Front Med 2025; 12: 1646412.

66 Vali Y, Lee J, Boursier J, Spijker R, Löffler J, Verheij J et al. Enhanced liver fibrosis test for the non-invasive diagnosis of fibrosis in patients with NAFLD: A systematic review and meta-analysis. J Hepatol 2020; 73: 252–262.

67 Martin EM, Chang J, González A, Genovese F. Circulating collagen type I fragments as specific biomarkers of cardiovascular outcome risk: Where are the opportunities? Matrix Biol 2025; 137: 19–32.

68 Tsimberidou AM, Fountzilas E, Nikanjam M, Kurzrock R. Review of Precision Cancer Medicine: Evolution of the Treatment Paradigm. Cancer Treat Rev 2020; 86: 102019.

