## Additional File 2 for "Urinary collagen type I degradation products as common fibrosis biomarkers in chronic diseases"

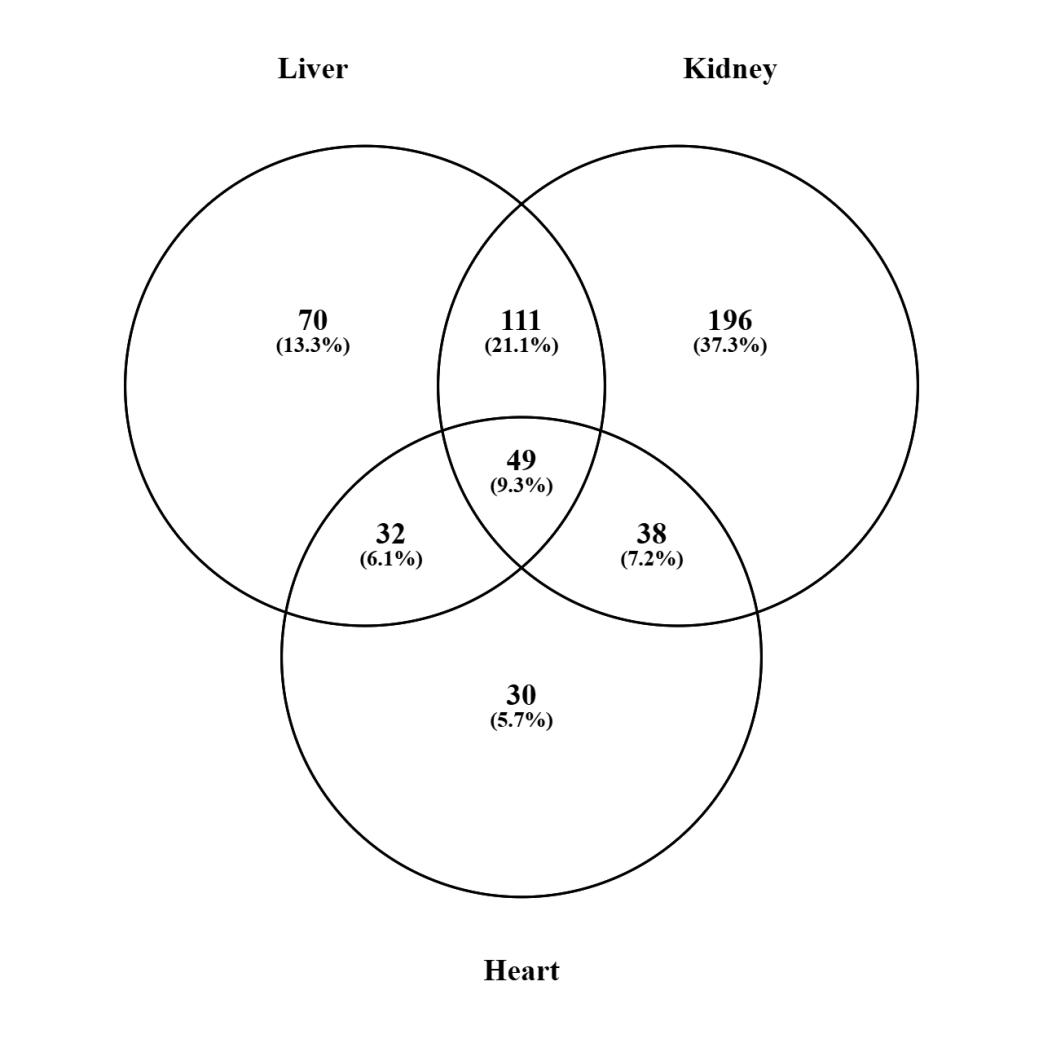

**Supplementary Figure 1. Venn diagram showing the overlap of fibrosis-associated COL1 peptides across different organ case-control comparisons.**

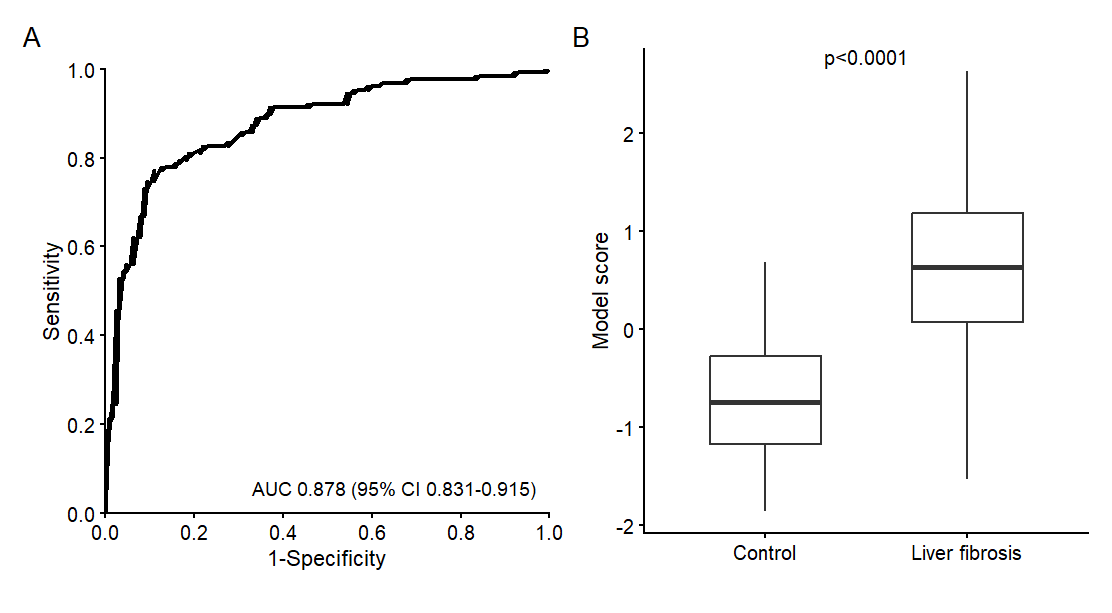

**Supplementary Figure 2. Performance of the collagen type I-based fibrosis model during cross-validation.** (A) Receiver operating characteristic (ROC) curve showing the performance of the 50-peptide model based on the cross-validation scores in the liver fibrosis training cohort, comprising patients with liver fibrosis (n=127) and matched controls (n=127). The model achieved an AUC of 0.878 (95% CI 0.831-0.915, p<0.0001). The optimal cut-off of >0.045 was determined using the Youden index and resulted in a sensitivity of 77.2% (95% CI 68.9-84.1%) and specificity of 89.0% (95% CI 82.2-93.8%). (B) Box plots showing the cross-validation model scores in patients with liver fibrosis and matched controls. Model scores were significantly higher in patients with liver fibrosis compared with controls (p<0.0001, Mann–Whitney test). Abbreviations: AUC: area under the receiver operating characteristic curve, CI: confidence interval.
